# Postgraduate education among family and community physicians in Brazil: the *Trajetórias MFC* project

**DOI:** 10.1101/19005744

**Authors:** Leonardo Ferreira Fontenelle, Stephani Vogt Rossi, Miguel Henrique Moraes de Oliveira, Diego José Brandão, Thiago Dias Sarti

**Author notes:** **Contribution** - Conceptualization: LFF (equal), DJB (supporting), TDS (equal) - Data curation: LFF - Formal analysis: LFF - Funding acquisition: none - Investigation: LFF (lead), SVR (supporting), MHMO (supporting) - Methodology: LFF (lead), DJB (supporting), TDS (supporting) - Project administration: LFF - Resources: none - Software: none - Supervision: none - Validation: none - Visualization: LFF - Writing-original draft: LFF (leading), SVR (supporting) - Writing-review & editing: SVR (supporting), MHMO (supporting), DJB (supporting), TDS (lead).

## Abstract

Neither primary health care or family and community medicine are recognized as knowledge areas in Brazil, for the purpose of postgraduate education (master’s, Ph.D.) or research. Our objective was to describe the postgraduate education trajectories of family and community physicians in Brazil. In this observational, exploratory study, we used data from SBMFC and SisCNRM to compile the list of physicians and community physicians, and then downloaded their curricula vitae from the Lattes Platform, verifying all data for consistency. A master’s degree was held by one in eight, and a Ph.D., by one in forty; most degrees were in collective health. Women (versus men) were less likely to hold master’s degrees, and even less likely to hold Ph.D. degrees. Professional (versus academic) master’s degrees and those in other areas (versus in medicine or collective health) were also associated with lower probability of obtaining a Ph.D. degree. Certified specialists (versus those with a medical residency) with a postgraduate degree were more likely to have earned it before becoming family and community physicians. We suggest that researchers in public health critically examine the relative benefits of different postgraduate trajectories for the professional performance of family and community physicians.

## Introduction

In the second half of the 20th century, family medicine differentiated from general practice as a result of the increased appreciation of the importance of primary health care, the psychosocial aspects of health and the complexity of causation.^1–3^ “General practitioners” in countries such as the United Kingdom and the Netherlands are expected to undergo postgraduate training (medical residency), and thus are family physicians by another name.^4–6^

In Brazil, emphasis on primary health care coincided with the fight for redemocratization, culminating in the establishment of a national health system called Sistema Único de Saúde (SUS; Unified Health System).^1,7–10^ The origin of family medicine in Brazil, however, was everything but unified. In the 1970s, movements called community medicine (*medicina comunitária*) and comprehensive medicine (*medicina integral*) merged into a medical specialty then called community general medicine (*medicina geral comunitária*).^1,7–9^ In 2000-2001, after SUS started expanding access to primary health care through the Family Health Strategy (then Family Health Program, conceived based on community general medicine and other inputs ^7,11,12^), community general medicine resolved its differences with family medicine, resulting in a medical specialty now called family and community medicine (*medicina de família e comunidade*, MFC).^1,8^ Preventive and social medicine (*medicina preventiva e social*) remains a separate specialty, despite the partially overlapping scope of practice.^1^

The concepts of “field of competence” and “core competence” ^13^ have been considered useful for understanding the interplay between the disciplines of primary health care and family and community medicine.^9^ Family and community medicine having its core competences does not preclude it from sharing primary health care as a field of competence, or vice-versa. Even though McWhinney and Freeman have argued family medicine to be an academic discipline separate from other medical disciplines,^2,14,15^ they also recognize distinctions among disciplines are sometimes more administrative and historical than epistemological.^2^

In Brazil, though, neither primary health care, nor family and community medicine are acknowledged as knowledge areas in the official “tree of knowledge” adopted by CAPES (the Federal Agency for Support and Evaluation of Graduate Education), CNPq (the National Council for Scientific and Technological Development) and FINEP (the Funding Authority for Studies and Projects). This means research grant applications and postgraduate programs (master’s and Ph.D.) must be registered in correlated basic knowledge areas, such as interdisciplinary, medicine or collective health, or in some subarea or specialty within those basic knowledge areas. In Latin America, “collective health” comprises (among other definitions ^16^) public health, epidemiology and humanities and social sciences in health.^17^

Accordingly, there are only a few postgraduate programs on primary health care or Family Health ^18–21^ (but no one on family and community medicine, that we know of) in Brazil, and university departments dedicated to primary health care and/or family and community medicine are exceedingly rare. Brazilian journals on primary health care and/or family and community medicine have little prestige, and thus a substantial part of relevant research is expected to be published in other, more general journals (this is not specific to Brazil ^22^). All this makes it very hard to gather a comprehensive understanding of the postgraduate education and research in primary health care and/or family and community medicine.

In this study, we hope to shed some light on the postgraduate education of primary health care professionals in Brazil by using specialization in family and community medicine as a way to identify physicians more likely to be involved with primary health care. Our objective was to explore their trajectories in postgraduate education (master’s and Ph.D.), describing the characteristics of their postgraduate degrees as well as correlating such degrees with characteristics of the physicians and their previous training.

## Methods

In this article, we report results from an observational, exploratory study, integrating secondary data from multiple sources: the *Trajetórias MFC* project.

### Data sources

We compiled the list of family and community physicians in Brazil from two sources, corresponding to the two modes of recognition as an specialist for physicians in Brazil.^23^ One mode is certification by the corresponding specialty association. Such specialty certificates are conferred to physicians with either experience or a medical residency who pass an exam occurring once or twice every year, since 2003.^3,8^ We obtained the list of certified family and community physicians, as of late November 2018, from Sociedade Brasileira de Medicina de Família e Comunidade (SBMFC, the Brazilian Society of Family and Community Medicine), corresponding to the first 24 editions of the specialist certification exam. For physicians having renewed their certification (which is not required), we kept only data on their first certification.

The other way of being recognized as an specialist is completing a medical residency. In late December 2018, we downloaded spreadsheets from SisCNRM (the information system for medical residency) on both family and community medicine and general community medicine (that is, from after and before the specialty was renamed) and merged the lists. Again, for the few physicians having completed a medical residency more than once, we kept only data on their first medical residency.

After obtaining the data from both sources, we merged the lists into a single list of family and community physicians, using both the name and the CPF (*Cadastro de Pessoas Físicas*) registry identification number. We verified this compiled list extensively for internal consistency, resorting to Web searches and to looking up in the Lattes Platform when additional information was necessary.

Lattes Platform is the Brazilian information system on science, technology, and innovation. Created in 1999, its curricula vitae (CVs) tend to be quite complete because they are used for decisions on research funding and on recruitment, promotion and tenure.^24^ Furthermore, CVs in the Lattes Platform tend to be honest because researchers are accountable for the information they provide. In example, their individual CVs are publicly available as Web pages and eXtensible Markup Language (XML) files.

After compiling the list of family and community physicians, we found their online CV Web pages using specially crafted uniform resource locators (URLs) including their CPF or looking up the Lattes Platform using the physicians’ names. Then, we obtained their Lattes Platform ID number by scraping these Web pages, and downloaded the corresponding CVs in XML format in late December 2018.

After extracting data on the postgraduate courses from the Lattes XML files, we hand checked each entry to make sure the data corresponded to actual postgraduate programs and each entry with a postgraduate program included the corresponding ID code. This hand checking included looking up the postgraduate programs in the Sucupira Platform (the Brazilian information system on postgraduate programs)^25^ and searching the Web for the monographs (titles are often included in the Lattes CV, and monographs are often publicly available). Then we obtained further data on the postgraduate programs from the Sucupira Platform, by scraping their web pages corresponding to each postgraduate program. For international programs, we imputed the code for the knowledge area and the mode of the master’s degree by hand, based on the program title.

### Variables

We inferred gender from the physician’s first name, using data from the 2010 Brazilian Census ^26^ cached in the genderBR package (version 1.1.0).^27^ First names were considered female if they had 50%+ probability of belonging to a woman, and male if otherwise; names appearing less than 20 times in the Census were not assigned any gender. The date when the Lattes CVs were last updated was retrieved from CVs themselves.

When the physician had both completed a medical residency and received a specialty certificate, we considered whichever came first as the mode of specialization. The year of specialization, master’s or Ph.D. completion was categorized in five-year periods, with the first period (up to 1998, the last year before inception of the Lattes Platform) having an open beginning. Likewise, the states where the specialization, master’s or Ph.D. took place were grouped into the five geographical regions (North, Northeast, Southeast, South and Central-West), with an extra category for international postgraduate programs (Supplemental Table ^28^ lists data for individual states). The knowledge areas were handled at the “basic knowledge area” level (that is, not at the “greater knowledge area” level, nor at the “sub-area” or “speciality” levels), and areas other than medicine or collective health were grouped in a catchall category (Supplemental Table ^28^ lists data for individual basic knowledge areas). Master’s degrees were also categorized according to their mode, that is, according to whether they were academic or professional. While both modes involve immersion in research, the academic master’s courses aim to produce researchers, and the professional ones aim to produce better professionals for outside academia.^29,30^ Professional Ph.D. courses are very recent, and we didn’t expect any degree to have been earned yet.

### Analysis

Categorical data were described with absolute and relative frequencies, and continuous data were described with medians and interquartile ranges (IQR).

Besides describing gender and characteristics of specialization and of master’s and Ph.D. degrees, we also described the frequency of master’s degrees among family and community physicians according to their gender and characteristics of specialization, as well as the frequency of Ph.D. degrees according to the same characteristics as well as to the characteristics of the master’s degree. Because the interpretation of the year of specialization depends on whether the specialization was through medical residency or through certification, year and mode of specialization were merged into a single variable for the purpose of describing the frequency of master’s and Ph.D. degrees.

The association of the before-mentioned explanatory variables with having earned a master’s degree (one response variable) or Ph.D. (another response variable) was expressed with the odds ratio (OR). While the prevalence ratio could be more intuitive, the OR better approximates the incidence density (rate) ratio.^31^ The OR were estimated through logistic regression both with a single and with multiple explanatory variables at a time, and were expressed as point estimates and 95% uncertainty interval. All regression coefficients (including the intercept) had weakly informative prior distributions,^32^ and inference was based on the No-U-Turn sampler.^33^ Year and mode of specialization were entered in such regression models separately along with an interaction term between them, but for ease of understanding the results were presented as if there was a single “year and mode of specialization” variable.

In the multivariable model for the Ph.D. degree, we opted for a hierarchical framework.^34^ The “distal level” included gender and characteristics of the specialization, and the “proximal level” included characteristics of the master’s. Variables in the distal level had their OR adjusted for other variables in the distal level, and variables in the proximal level had their OR adjusted for all other variables in the model, from both levels.

Time from specialization to master’s and Ph.D. degrees, and between the two later ones, was described both in general and according to the mode of specialization. We also examined the distribution of the knowledge areas of the master’s degrees according to the mode of specialization, and the distribution of knowledge areas of Ph.D. degrees according to the mode of specialization and the knowledge area of the master’s. We did not examine the distribution of the knowledge area of the Ph.D. degree according to the mode of the master’s, because too few family and community physicians with a professional master’s earned a Ph.D.. When describing the distribution of the knowledge area of the Ph.D. according to the knowledge area of the master’s, we opted for an alluvial diagram.

The data were analyzed using R (version 3.6.1).^35^ While data verification by hand involved spreadsheet applications from office suites, most data processing (including the tabulation of frequencies) was done within R itself, using packages centered around the concept of “tidy data”,^36^ as well as packages specific to Brazilian data.^27,37^ For regression modelling we used brms (version 2.10.0),^38^ which builds on the Stan probabilistic programming language ^39^ through rstan (version 2.19.2).^40^ The alluvial diagram was made with ggalluvial (version 0.10.0).^41^

### Ethics

While most of the data sources we used are publicly available, there were two privacy issues. First, the spreadsheet SBMFC provided us with the complete list of certificates (including the CPF number of most) is not publicly available as such. Second, while all data we obtained from the Lattes Platform are publicly available, they lied behind technology clearly intended no hinder data mining. Thus, before initiating the data collection we obtained approval by the research ethics committee of Universidade Vila Velha (certificate 02957118.2.0000.5064). Because our data is personally identifiable, it will be available to other researchers only if they present an ethically-approved research project with an analysis plan ^42^.

## Results

The first 24 editions of the specialist certification exam resulted in the emission of 2816 certificates, amounting to 2795 unique physicians. Furthermore, 3959 medical residencies were completed (936 in general community medicine and 3023 after it was renamed family and community medicine), amounting to 3957 unique physicians. Because 514 physicians both concluded a medical residency and were certified, there were 6238 unique family and community physicians. Of these, 4065 (65.2%) had a CV in the Lattes Platform. Median time since these CVs were last updated was 1 year, with an IQR of 0–4 years and a maximum of 18 years.

Most family and community physicians (3563, 58.3%) were female, and the most common mode of specialization was medical residency (3917, 62.8%) (Table 1). The number of new family and community physicians clearly increased over time, with the largest increases being from 1999–2003 to 2004–2008 and from 2009–2013 to 2014–2018. Specialization in family and community medicine was concentrated in the Southeast and South regions.

**Table 1:** Family and community physicians in Brazil, December 2018

| **Characteristic** | **n** | **%** |
| --- | --- | --- |
| **Gender** |  |  |
| Female | 3563 | 58.3% |
| Male | 2546 | 41.7% |
| **Mode of specialization** |  |  |
| Certification | 2321 | 37.2% |
| Medical residency | 3917 | 62.8% |
| **Year of specialization** |  |  |
| 2014–2018 | 2364 | 37.9% |
| 2009–2013 | 1573 | 25.2% |
| 2004–2008 | 1368 | 21.9% |
| 1999–2003 | 370 | 5.9% |
| 1981–1998 | 563 | 0 |
| **Region of specialization** |  |  |
| North | 314 | 5.1% |
| Northeast | 825 | 13.3% |
| Southeast | 2794 | 45.1% |
| South | 1943 | 31.4% |
| Central-West | 317 | 5.1% |

A master’s degree was obtained by 747 (12.0%) family and community physicians (Table 2), with 554 (74.2%) degrees being academic and the other 193 (25.8%) being professional. The number of new master’s degrees clearly increased with time, especially from 2004–2008 to 2009–2013; from 2009–2013 to 2014–2018, the increase was restricted to professional master’s degrees. As for specialization, master’s degrees were concentrated in the Southeast and South regions, both for academic and professional degrees. Master’s degrees in collective health (351, 47.0%) were twice as frequent as those in medicine (170, 22.8%), with other frequent knowledge areas being interdisciplinary (84, 11.2%), teaching (30, 4.0%) and education (24, 3.2%). Professional degrees accounted for 104 (29.6%) of the master’s degrees in collective health, but only 27 (15.9%) of those in medicine. The postgraduate programs were spread across 141 institutions, with only eight of them having conferred a master’s degree to at least 20 family and community physicians. Most family and community physicians obtained their master’s degrees from Universidade Federal do Rio Grande do Sul (UFRGS; Federal University of Rio Grande do Sul), Fundação Oswaldo Cruz (Fiocruz; Oswaldo Cruz Foundation) or Universidade de São Paulo (USP; University of São Paulo); more information on Supplemental Table ^28^.

**Table 2:**
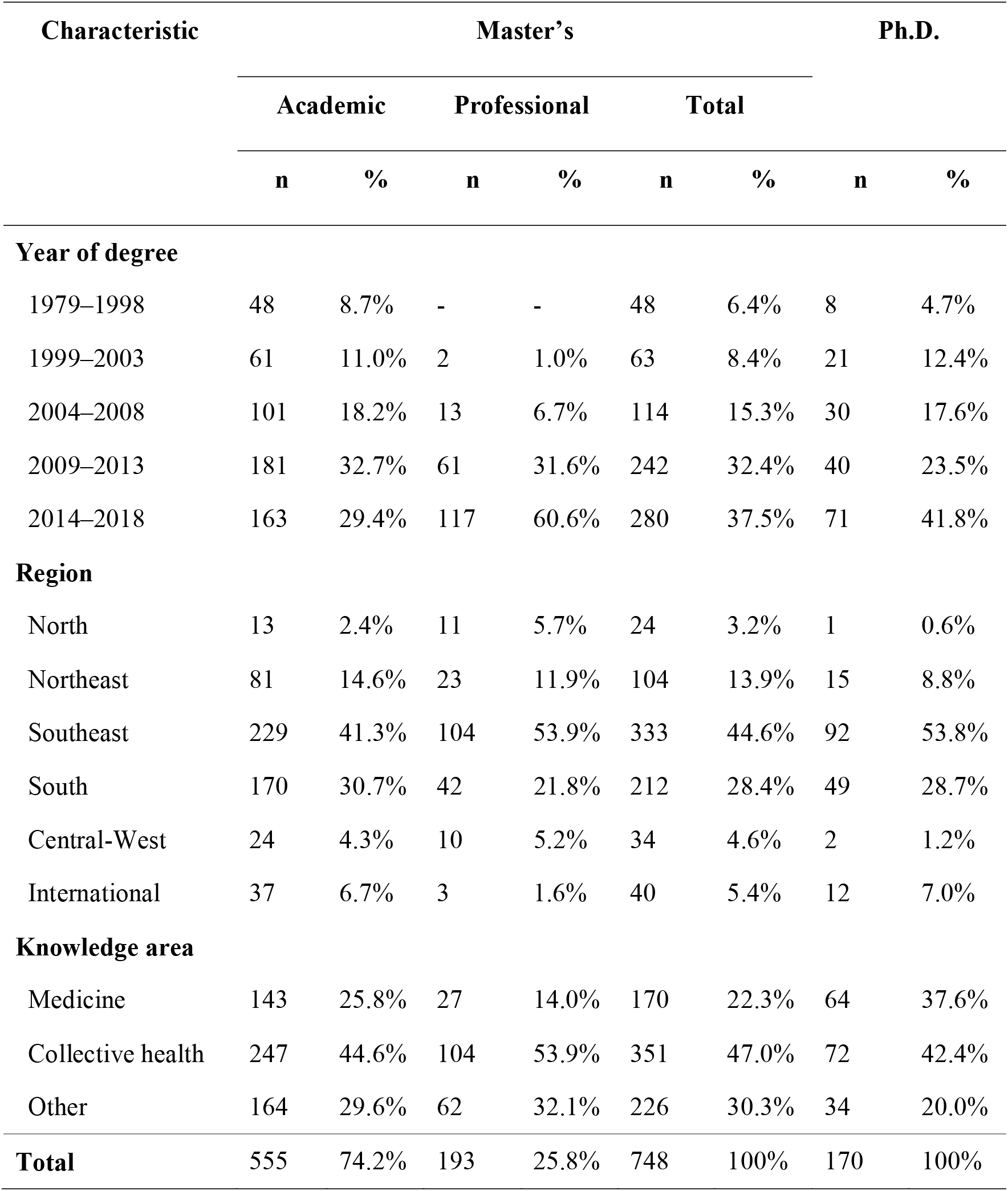
Master’s and Ph.D. degrees of family and community physicians in Brazil, December 2018

A Ph.D. degree was held by 170 (2.7%) family and community physicians (Table 2). There were no professional Ph.D. degrees. As with master’s degrees, there was an increase in new Ph.D. degrees over time, and most degrees were obtained in the Southeast and South regions of Brazil. Contrary to master’s degrees, Ph.D. degrees in medicine (64, 38%) were almost as common as those in collective health (72, 42%). The postgraduate programs were spread across 40 institutions, with only two of them (USP and UFRGS) having conferred more than 20 degrees; more information on Supplemental Table ^28^.

Some groups of family and community physicians were less likely to hold a master’s or Ph.D. degree, even after adjusting for other characteristics (Table 3, Table 4). Women were less likely to hold a master’s degree than their male colleagues, and only half as likely to hold a Ph.D.. Family and community physicians with a professional master’s degree were □ as likely to earn a Ph.D. degree as those with an academic master’s degree. A master’s degree in other knowledge area also implicated lower likelihood of earning a Ph.D. than a master’s in medicine or collective health.

**Table 3:** Characteristics associated with holding a master’s degree among family and community physicians in Brazil, 2018

| Characteristic | Frequency |  | Raw model |  | Adjusted model |  |
| --- | --- | --- | --- | --- | --- | --- |
|  | n | % | OR | 95% UI | OR | 95% UI |
| **Gender** |  |  |  |  |  |  |
| Female | 384 | 10.8% | 1 | (reference) | 1 | (reference) |
| Male | 353 | 13.9% | 1.33 | 1.15–1.54 | 1.24 | 1.07–1.45 |
| **Mode and year of specialization \*** |  |  |  |  |  |  |
| Certification, 2014–2018 | 94 | 13.4% | 1 | (reference) | 1 | (reference) |
| Certification, 2009–2013 | 70 | 8.8% | 0.64 | 0.48–0.84 | 0.64 | 0.48–0.84 |
| Certification, 2004–2008 | 137 | 17.1% | 1.35 | 1.08–1.72 | 1.37 | 1.08–1.76 |
| Certification, 1999–2003 | 7 | 26.9% | 2.33 | 1.07–4.72 | 2.53 | 1.12–5.11 |
| Residency, 2014–2018 | 71 | 4.3% | 0.30 | 0.23–0.38 | 0.31 | 0.23–0.40 |
| Residency, 2009–2013 | 114 | 14.6% | 1.11 | 0.87–1.42 | 1.17 | 0.90–1.49 |
| Residency, 2004–2008 | 118 | 21.0% | 1.72 | 1.32–2.19 | 1.75 | 1.34–2.27 |
| Residency, 1999–2003 | 64 | 18.6% | 1.49 | 1.11–2.00 | 1.60 | 1.19–2.14 |
| Residency, 1981–1998 | 72 | 12.8% | 0.95 | 0.73–1.25 | 0.97 | 0.73–1.30 |
| **Region of specialization** |  |  |  |  |  |  |
| North | 32 | 10.2% | 0.92 | 0.61–1.35 | 1.27 | 0.81–1.89 |
| Northeast | 104 | 12.6% | 1.17 | 0.92–1.48 | 1.19 | 0.93–1.51 |
| Southeast | 306 | 11.0% | 1 | (reference) | 1 | (reference) |
| South | 260 | 13.4% | 1.25 | 1.05–1.49 | 1.02 | 0.84–1.22 |
| Central-West | 36 | 11.4% | 1.03 | 0.72–1.44 | 1.11 | 0.75–1.61 |

OR, odds ratio. UI, uncertainty interval. \* Specialist certification certification in family and community medicine began in 2003, whence few physicians were certified in 1999–2003, and none were certified before 1999.

**Table 4:**
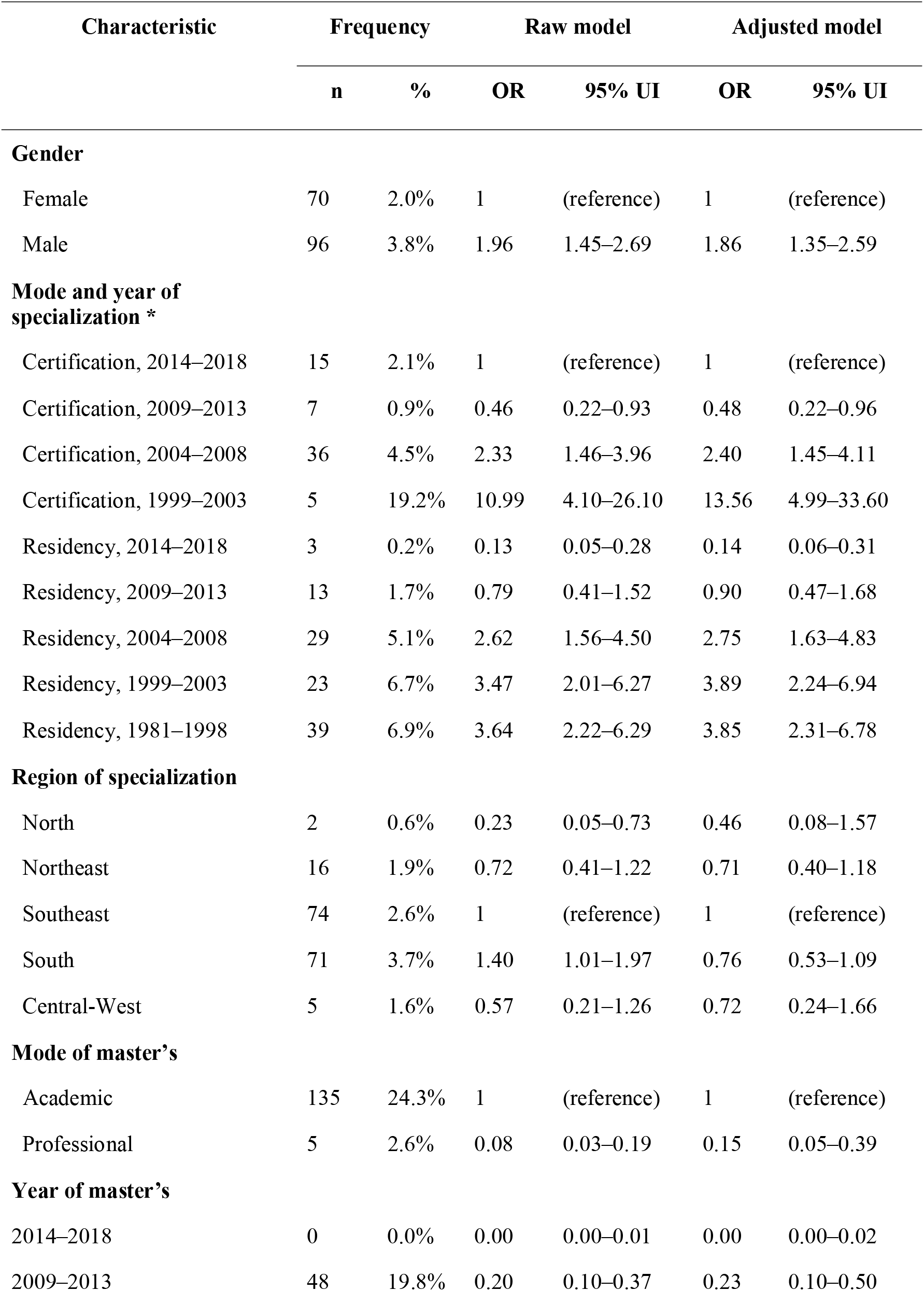
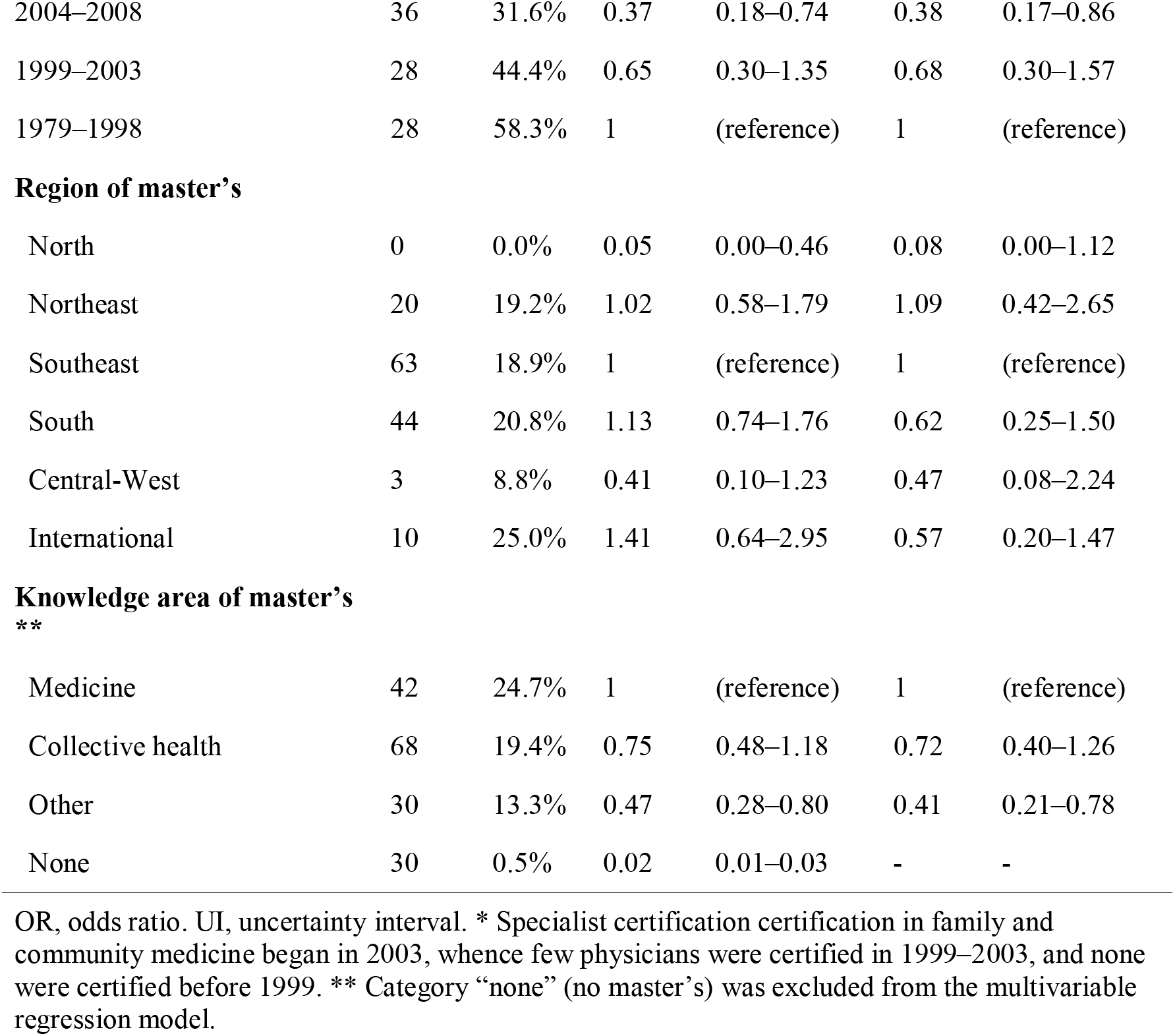
Characteristics associated with holding a Ph.D. degree among family and community physicians in Brazil, 2018

| Characteristic | Frequency |  | Raw model |  | Adjusted model |  |
| --- | --- | --- | --- | --- | --- | --- |
|  | n | % | OR | 95% UI | OR | 95% UI |
| **Gender** |  |  |  |  |  |  |
| Female | 70 | 2.0% | 1 | (reference) | 1 | (reference) |
| Male | 96 | 3.8% | 1.96 | 1.45–2.69 | 1.86 | 1.35–2.59 |
| **Mode and year of specialization \*** |  |  |  |  |  |  |
| Certification, 2014–2018 | 15 | 2.1% | 1 | (reference) | 1 | (reference) |
| Certification, 2009–2013 | 7 | 0.9% | 0.46 | 0.22–0.93 | 0.48 | 0.22–0.96 |
| Certification, 2004–2008 | 36 | 4.5% | 2.33 | 1.46–3.96 | 2.40 | 1.45–4.11 |
| Certification, 1999–2003 | 5 | 19.2% | 10.99 | 4.10–26.10 | 13.56 | 4.99–33.60 |
| Residency, 2014–2018 | 3 | 0.2% | 0.13 | 0.05–0.28 | 0.14 | 0.06–0.31 |
| Residency, 2009–2013 | 13 | 1.7% | 0.79 | 0.41–1.52 | 0.90 | 0.47–1.68 |
| Residency, 2004–2008 | 29 | 5.1% | 2.62 | 1.56–4.50 | 2.75 | 1.63–4.83 |
| Residency, 1999–2003 | 23 | 6.7% | 3.47 | 2.01–6.27 | 3.89 | 2.24–6.94 |
| Residency, 1981–1998 | 39 | 6.9% | 3.64 | 2.22–6.29 | 3.85 | 2.31–6.78 |
| **Region of specialization** |  |  |  |  |  |  |
| North | 2 | 0.6% | 0.23 | 0.05–0.73 | 0.46 | 0.08–1.57 |
| Northeast | 16 | 1.9% | 0.72 | 0.41–1.22 | 0.71 | 0.40–1.18 |
| Southeast | 74 | 2.6% | 1 | (reference) | 1 | (reference) |
| South | 71 | 3.7% | 1.40 | 1.01–1.97 | 0.76 | 0.53–1.09 |
| Central-West | 5 | 1.6% | 0.57 | 0.21–1.26 | 0.72 | 0.24–1.66 |
| **Mode of master's** |  |  |  |  |  |  |
| Academic | 135 | 24.3% | 1 | (reference) | 1 | (reference) |
| Professional | 5 | 2.6% | 0.08 | 0.03–0.19 | 0.15 | 0.05–0.39 |
| **Year of master's** |  |  |  |  |  |  |
| 2014–2018 | 0 | 0.0% | 0.00 | 0.00–0.01 | 0.00 | 0.00–0.02 |
| 2009–2013 | 48 | 19.8% | 0.20 | 0.10–0.37 | 0.23 | 0.10–0.50 |
| 2004–2008 | 36 | 31.6% | 0.37 | 0.18–0.74 | 0.38 | 0.17–0.86 |
| 1999–2003 | 28 | 44.4% | 0.65 | 0.30–1.35 | 0.68 | 0.30–1.57 |
| 1979–1998 | 28 | 58.3% | 1 | (reference) | 1 | (reference) |
| **Region of master's** |  |  |  |  |  |  |
| North | 0 | 0.0% | 0.05 | 0.00–0.46 | 0.08 | 0.00–1.12 |
| Northeast | 20 | 19.2% | 1.02 | 0.58–1.79 | 1.09 | 0.42–2.65 |
| Southeast | 63 | 18.9% | 1 | (reference) | 1 | (reference) |
| South | 44 | 20.8% | 1.13 | 0.74–1.76 | 0.62 | 0.25–1.50 |
| Central-West | 3 | 8.8% | 0.41 | 0.10–1.23 | 0.47 | 0.08–2.24 |
| International | 10 | 25.0% | 1.41 | 0.64–2.95 | 0.57 | 0.20–1.47 |
| Medicine | 42 | 24.7% | 1 | (reference) | 1 | (reference) |
| Collective health | 68 | 19.4% | 0.75 | 0.48–1.18 | 0.72 | 0.40–1.26 |
| Other | 30 | 13.3% | 0.47 | 0.28–0.80 | 0.41 | 0.21–0.78 |
| None | 30 | 0.5% | 0.02 | 0.01–0.03 | - | - |

OR, odds ratio. UI, uncertainty interval. \* Specialist certification certification in family and community medicine began in 2003, whence few physicians were certified in 1999–2003, and none were certified before 1999. \*\* Category “none” (no master's) was excluded from the multivariable regression model.

Among those earning a master’s degree, median time from specialization to master’s was 4 years (IQR, 0–6): 5 years (IQR, 3–8) for family and community physicians specializing through medical residency, and 1 year (IQR, −3 to 4) for those specializing through certification. In other words, earning a master’s degree came before specialization for 21.8% family and community physicians: 9.1% for those specializing through medical residency, and 39.9% for those specializing through certification. The proportion of master’s degrees in each of the knowledge areas (medicine, collective health, other) was very similar across the specialization modes (certification or medical residency) (data not shown).

Likewise, median time from specialization to Ph.D. degree was 9 years (IQR, 3.25– 12): 10 years (IQR, 7–14) for family and community physicians specializing through medical residency, and 2 years (IQR, −2.5 to 9) for those specializing through certification. In other words, earning a Ph.D. degree came before specialization for 16% family and community physicians: 6% for those specializing through medical residency, and 33% for those specializing through certification. As with the master’s degrees, the proportion of Ph.D. degrees in each of the knowledge areas (medicine, collective health, other) was very similar across the specialization modes (certification or medical residency) (data not shown).

Finally, median time from master’s to Ph.D. degree was 4 years (IQR, 0–6). Most family and community physicians earning a Ph.D. in collective health (50, 69%) also earned a master’s in the same knowledge area (Graph 1). On the other hand, half of those earning a Ph.D. in medicine earned a master’s in collective health (11, 17%) or no master’s at all (i.e., direct Ph.D.; 19, 30%). Most direct Ph.D. degrees (16, 53%), in medicine or otherwise, were conferred by USP.

**Graph 1:**
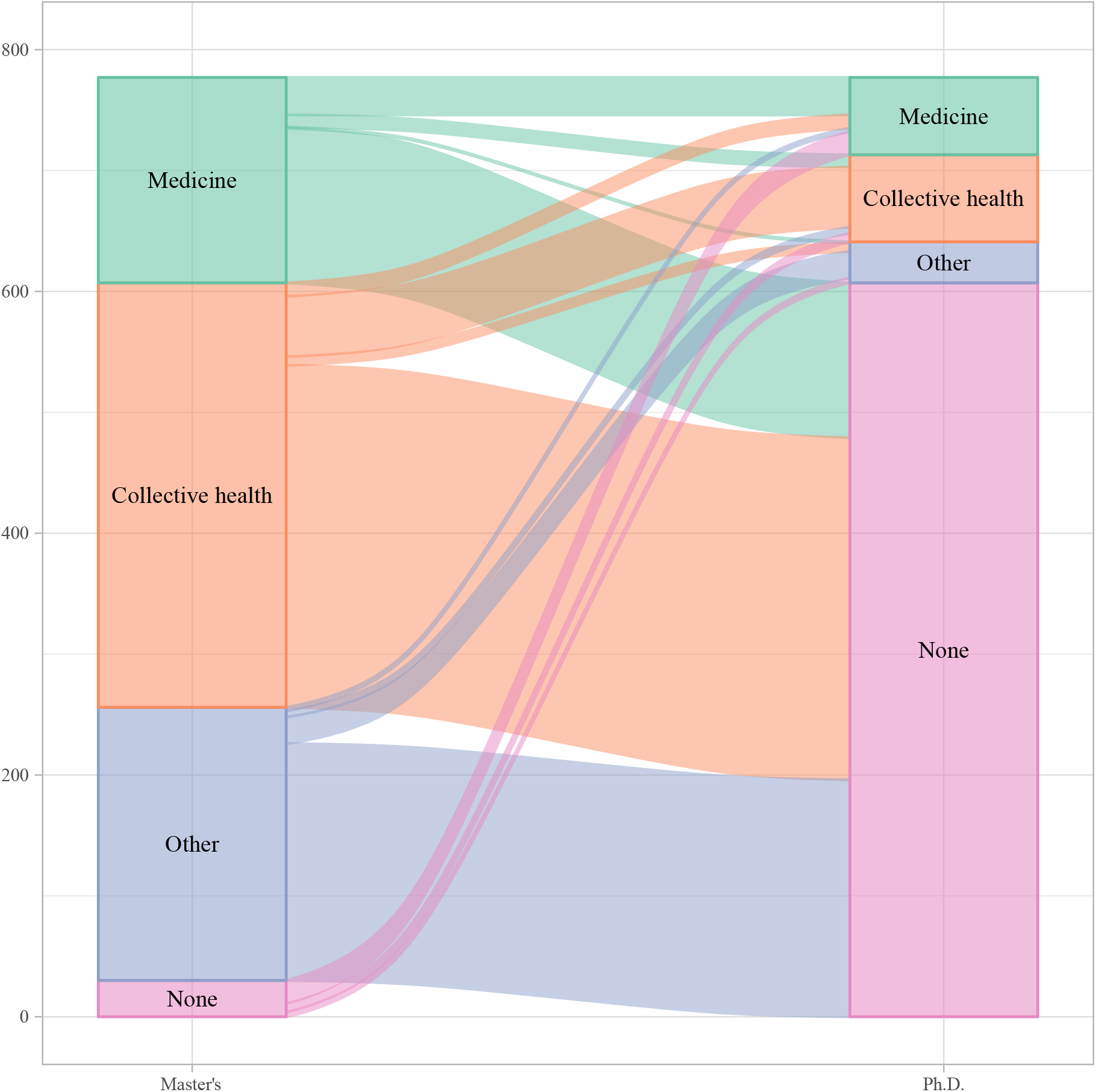
Alluvial diagram of knowledge areas for master’s and Ph.D. degrees of family and community physicians in Brazil, 2018

## Discussion

Our findings indicate one in eight family and community physicians have earned a master’s degree, and little more than one in forty have earned a Ph.D.. The number of new degrees is increasing over time, and most of the degrees are in collective health (not medicine) and were obtained in the Southeast and South regions. Gender is associated with the probability of obtaining a master’s degree and, together with mode and knowledge area of the master’s, also with that of obtaining a Ph.D. degree. In comparison to family and community physicians specializing through medical residency, those specializing through certification were more likely to already have earned a master’s and/or Ph.D. degree (instead of earning after specialization), but just as likely to have earned their degree in medicine, collective health or other knowledge areas.

Professional master’s programs are responsible for most of the recent increase in the number of master’s degrees, and might be attracting graduate students who would otherwise opt for academic programs. The Northeast region, with its RENASF network of institutions offering a professional master’s program focused on the Family Health Strategy,^19,20^ accounts for part of this increase, as does the state of Rio de Janeiro, whose capital city recently invested heavily in the expansion of the Family Health Strategy and incentivized health professionals to earn master’s degrees;^21^ but the increase in professional master’s degrees was not restricted to these states (data not shown). Even though the ProfSaúde program ^18^ is expected to contribute to this increase, it does not account for our findings, because its first students graduated after we obtained our data.

Brazil has been promoting professional master’s on Family Health alongside other qualification initiatives, such as residencies, short postgraduate courses and academic master’s.^21,30^ While one would expect any qualification to contribute something, responsible public policies for human resources in health depend on critically examining the relative benefits of the multiple possible postgraduate trajectories. In example, some qualification initiatives might add little to the performance of professionals who are already sufficiently qualified, or who are not qualified enough to benefit from said initiatives. Furthermore, for better or for worse, a professional master’s degree should increase employability in academia, thereby increasing workforce turnover in the Family Health Strategy while contributing to the education of the new generation of health professionals.

This debate is made even more timely by the advent of professional Ph.D. programs. Graduates from professional master’s courses seem to be less likely to earn Ph.D. degrees, even after adjusting for potential confounders, such as when was the master’s concluded. Because graduates from professional programs are expected to be outside academia, they might not find value in academic Ph.D. courses. It remains to be seen if professional Ph.D. programs are needed to fill this niche, or if simply there’s not much use for any Ph.D. outside academia. Meanwhile, physicians are still not required to complete medical residency (or otherwise be certified) before working in primary care in Brazil, and posts in medical residencies lag behind the annual number of newly graduated physicians (the same applies for nurses).^43^ Another major finding is that most family and community physicians hold master’s and Ph.D. degrees in collective health, not medicine. This phenomenon is more common for master’s than for Ph.D. degrees, and for professional than for academic degrees, but it occurs in both levels and both modes of postgraduate programs. We can’t say this came as a surprise: of the authors who are family and community physicians, all three hold a Ph.D. in a subarea of collective health. In our experience, not only has collective health devoted substantial interest to primary health care as a public policy, but also the Brazilian medical community hasn’t devoted much interest to primary care as a locus of healthcare delivery. In example, searching for (“family medicine” OR “family and community medicine”) in the SciELO Brazil collection, one of the top five journals is on medical education and the other four are journals on collective health. This suggests most research on primary health care in Brazil to be on health policy, service management and health promotion, as well as medical education, but not so much on clinical care.

Most medical residencies, specialist certifications, masters’ and Ph.D. happened in the Southeast and South regions. The reason is twofold: these are two of the most populous regions (together with the Northeast region), and are the most economically developed ones. These facts also reflect in the overall distribution of physicians in Brazil.^44^ Interestingly, our data do not support a higher probability of obtaining a master’s or Ph.D. degree for family and community physicians specializing or obtaining a master’s degree in the Southeast and South regions. This is not to say our data support equity in access to postgraduate programs: there simply are too few family and community physicians outside the Southeast and South regions for us to make precise estimates. As the saying goes, “absence of evidence is not evidence of absence [of effect]”.^45^

On the other hand, the association of gender and postgraduate degrees was very clear: female family and community physicians are less likely to obtain master’s degrees than their male colleagues, and even less so for Ph.D. degrees. This should not be interpreted as gender having a direct effect on educational achievement: our study was exploratory, and inclusion of gender as an explanatory variable was motivated mostly by the SAGER guidelines.^46^ Rather, this finding should taken as justification for further studies, aimed at a more proper explanation for this correlation. Such explanation might have more to do with family and community medicine than with the Brazilian society at large, because half the master’s and Ph.D. graduates in Brazil are women.^47^ Besides the ethical relevance of elucidating and preferably removing any gendered barriers to postgraduate education, the issue has special relevance to the discipline because most family and community physicians are women.

In interpreting our findings, one must keep in mind we depended on administrative data. Because data from Conselho Federal de Medicina (CFM; the Federal Board of Medicine) is not available to others, we could not perform record linkage to identify who had retired, emigrated or deceased. The National Registration of Specialists was expected to provide easy access to an authoritative list of physicians in any medical specialty, but unfortunately it has not been maintained as prescribed by the More Doctors Law.^48^ Consequently, the number of family and community physicians is surely overestimated, even if we expect this overestimation to be minor, because family and community medicine is a fairly young specialty in Brazil.^44^ In 2018, Augusto *et al*.^23^ (using the same methods as us) estimated there would be 5.438 family and community physicians in Brazil, 276 (5%) more than the 5.162 found by Scheffer *et al*.,^44^ who had access to CFM data. Another limitation of our study was that, as in Augusto *et al*.,^23^ we could not include the family and community physicians who completed their medical residencies in the 1970s. SisCNRM records for family and community medicine (then community general medicine) begin at 1981, when the specialty was recognized; we can only hope family and community physicians from the 1970s eventually took the exams and were certified after 2003.

Our data had limitations with regard to postgraduate programs, as well. Postgraduate degrees will probably have been underreported to some extent, even though postgraduate students are incentivized to have a CV in the Lattes Platform and entering data on a postgraduate degree is simple enough. Furthermore, there surely was some information error in the reported degrees, but we hope to have cleaned most of those errors during our verification. One potential issue is that we verified the data using current information on the postgraduate programs, not information from when the degrees were obtained; but much of the information is not expected to change in any significant way, and possible changes in the knowledge subarea or specialty of the postgraduate programs would not matter for our analysis, which were done at the level of basic knowledge area. Finally, because the Lattes Platform was launched in 1999, master’s and Ph.D. degrees earned before then are expected to be underreported, even if this underreporting is expected to occur among those who don’t make much use of the degrees.

In conclusion, family and community physicians increasingly earn academic and professional master’s and Ph.D. degrees, with an emphasis on collective health, even though women seemingly face barriers to advance their education. The consequences of different postgraduate trajectories on professional performance and on primary health care research are unknown. We suggest it would be easier to gain a wider view of postgraduate education of primary health care professionals and/or on primary health care if the National Registration of Specialists came back online, and if primary health care and/or family and community medicine were recognized as knowledge areas.

## Data Availability

Because our data is personally identifiable, it will be available to other researchers only if they present an ethically-approved research project with an analysis plan.

https://doi.org/10.5281/zenodo.3376310

https://doi.org/10.5281/zenodo.3381576

## Acknowledgments

MHMO receives a scientific initiation scholarship from UVV.

